# Pallidal and motor cortical interactions determine gait initiation dynamics in Parkinson’s disease

**DOI:** 10.1101/2024.12.12.24318797

**Authors:** Jessica E. Bath, Kenneth H. Louie, Hamid Fekri Azgomi, Jannine P. Balakid, Kara N. Presbrey, Jacob H. Marks, Thomas A. Wozny, Doris D. Wang

## Abstract

Gait initiation is a fundamental human task, requiring one or more anticipatory postural adjustments (APA) prior to stepping. Deviations in amplitude and timing of APAs exist in Parkinson’s disease (PD), causing dysfunctional postural control which increases the risk of falls. The motor cortex and basal ganglia have been implicated in the regulation of postural control, however, their dynamics during gait initiation, relationship to APA metrics, and response to pharmacotherapy such as levodopa are unknown.

To address these questions, we streamed electrocorticography (ECoG) potentials from the premotor and primary motor cortices, as well as local field potentials (LFPs) from the globus pallidus in five people with PD exhibiting gait and balance dysfunction during a cued gait initiation task. Amplitude and timing of APA were evaluated with force plates and synchronized to the neural data. Subjects performed gait initiation trials under ON and LOW levodopa conditions to assess effects of medication on APA metrics and underlying neural dynamics.

All subjects demonstrated pallidal and cortical oscillatory changes during different phases of gait initiation. Grouped analysis revealed that from quiet standing to the first foot step, pallidal beta power showed stepwise decrease and broadband gamma power increases, whereas cortical potentials showed low frequency (theta, alpha, beta) power decrease during gait initiation, regardless of medication state. The pallidum and motor cortices also became increasingly coherent during gait initiation compared to quiet standing prior to APA onset. Using linear mixed models, we found that while pallidal gamma powers are predictive of APA scaling, pallidal-cortical coherence (theta, alpha, beta) and premotor-M1 gamma coherence are predictive of APA timing.

Our study is the first detailed characterization of basal-ganglia cortical circuit dynamics during human gait initiation. We identified significant pallidal motor cortical power and coherence changes that underlie the amplitude and timing of APA which appear to be independent of medication states of the study subjects. Our results provide evidence for a model where synchronized premotor and motor cortical activities transiently couple with the globus pallidus to regulate the timing of postural responses, and local pallidal activity regulate the amplitude of postural changes during gait initiation. It suggests that abnormal pallidal outflow and synchronization between the pallidum and motor cortices may be a pathophysiological mechanism underlying disordered postural response in Parkinson’s disease.

## Introduction

Gait initiation is a fundamental, bipedal motor task requiring precise coordination of the limbs under shifting balance conditions for safe and effective performance. Successful gait initiation requires an anticipatory postural adjustment (APA), consisting of muscle activations immediately preceding stepping to transfer the body’s center of mass forward over the stance foot while shifting the center of pressure to the stepping foot.^1,2^ While APAs are generally thought to be stereotyped in healthy individuals, deviations in APA timing and amplitude are often found in patients with Parkinson’s disease (PD)^1,3^ and can reflect postural instability.

Postural instability exists in approximately 20% of people with PD at disease onset and increases in prevalence and severity with disease duration.^4,5^ The symptom has been associated with negative sequelae including falls, freezing of gait (FoG), and decreased independence and quality of life.^5–8^ Existing interventions such as medication, deep brain stimulation (DBS), and rehabilitation, do not reliably produce meaningful or long-term improvements in postural responses and can even exacerbate symptoms.^9–12^

A primary reason for the difficulty in effectively treating postural instability in PD could be the inherent complexity of the neural circuits underlying postural control. It is theorized that PD’s pathology at the basal ganglia alters the timing relationship between the postural response and the step execution components of effective gait initiation, thus contributing to dysfunction.^13^ Evidence for the neural control of gait initiation in humans and issues in PD remains limited, however. Noninvasive electroencephalography (EEG) in healthy adults found that normal gait initiation is associated with sensorimotor mu (8-13 Hz) and beta (16-30 Hz) cortical event-related desynchronizations (ERDs) prior to movement, followed by event related synchronizations during APA and stepping at mu, beta, and gamma (30-40 Hz) frequencies.^14^ These cortical areas were also implicated during gait initiation failure, with decreased low beta ERDs and increased mu ERDs observed in control subjects and elevated high beta (21-38 Hz) power in people with PD.^15,16^ No study to date has directly recorded from the basal ganglia during gait initiation. High frequency stimulation of the subthalamic nucleus (STN) and globus pallidus internus (GPi) with DBS electrodes were shown to alter APAs (positively, on APA amplitudes and restoration of physiological muscle synergies,^17^ or negatively, through delayed and ineffective compensatory stepping),^18^ indicating the importance of basal ganglia-thalamo-cortical circuits in coordinating this task.^19^

Given the limited understanding of the neurophysiology of postural control in humans, we sought to elucidate the cortical-basal ganglia circuit dynamics that underlie gait initiation and APA dysfunction in PD patients. We hypothesized that prior to the taking the first step from a standing position, the premotor cortex may produce a motor program that will engage the primary motor cortex (M1) for execution. This planned postural response also requires the dynamic coupling between the cortex and basal ganglia for refinement and the scaling before the first step. To this end, we streamed ambulatory local field potentials (LFP) from the GPi and globus pallidus externus (GPe), and cortical electrocorticography (ECoG) potentials from M1 and premotor cortex (PMC) in PD patients undergoing a cued gait initiation task under different dopaminergic medication states using an investigational bidirectional DBS device. We computed pallidal and motor cortical oscillatory power and coherence changes within and between the basal ganglia and cortex throughout the gait initiation task. We varied medication states to evaluate effects on APA and changes in network communication associated with the amplitude, scaling, and timing components of APAs.

We found that gait initiation is associated with progressive pallidal beta power decreases and broadband gamma increases, as well as cortical lower frequency power decreases, regardless of medication state. The motor cortical regions and pallidum also become coupled across multiple frequency bands during gait initiation compared to quiet standing. Linear mixed models (LMMs) suggest that while pallidal gamma powers are predictive of APA amplitudes, intra-cortical gamma coherence and pallidal-cortical low frequency coherence are predictive of APA timing. Our results highlight cortical-pallidal network dynamics in regulating gait initiation and provide evidence of abnormal network synchronization associated with disordered gait initiation in PD.

## Materials and Methods

### Study participants

#### Subjects

Five subjects with idiopathic PD evaluated for DBS surgery were enrolled in the clinical trial (ClinicalTrials.gov ID: NCT-03582891) at the University of California, San Francisco (Table 1). Study inclusion criteria included: motor fluctuations (>25% improvement between medication states), OFF medication gait disturbances, walk without assistive device, <4 falls/month, and Montreal Cognitive Assessment score (MoCA) ≥ 21. Study exclusion criteria included: ON medication FoG, Hoehn-Yahr stages ≥3, atypical PD diagnosis, chronic medical conditions affecting gait and balance or ability to perform activity. All subjects provided written informed consent according to the Declaration of Helsinki and the study was administered under institutional review board approval.

**Table 1:**
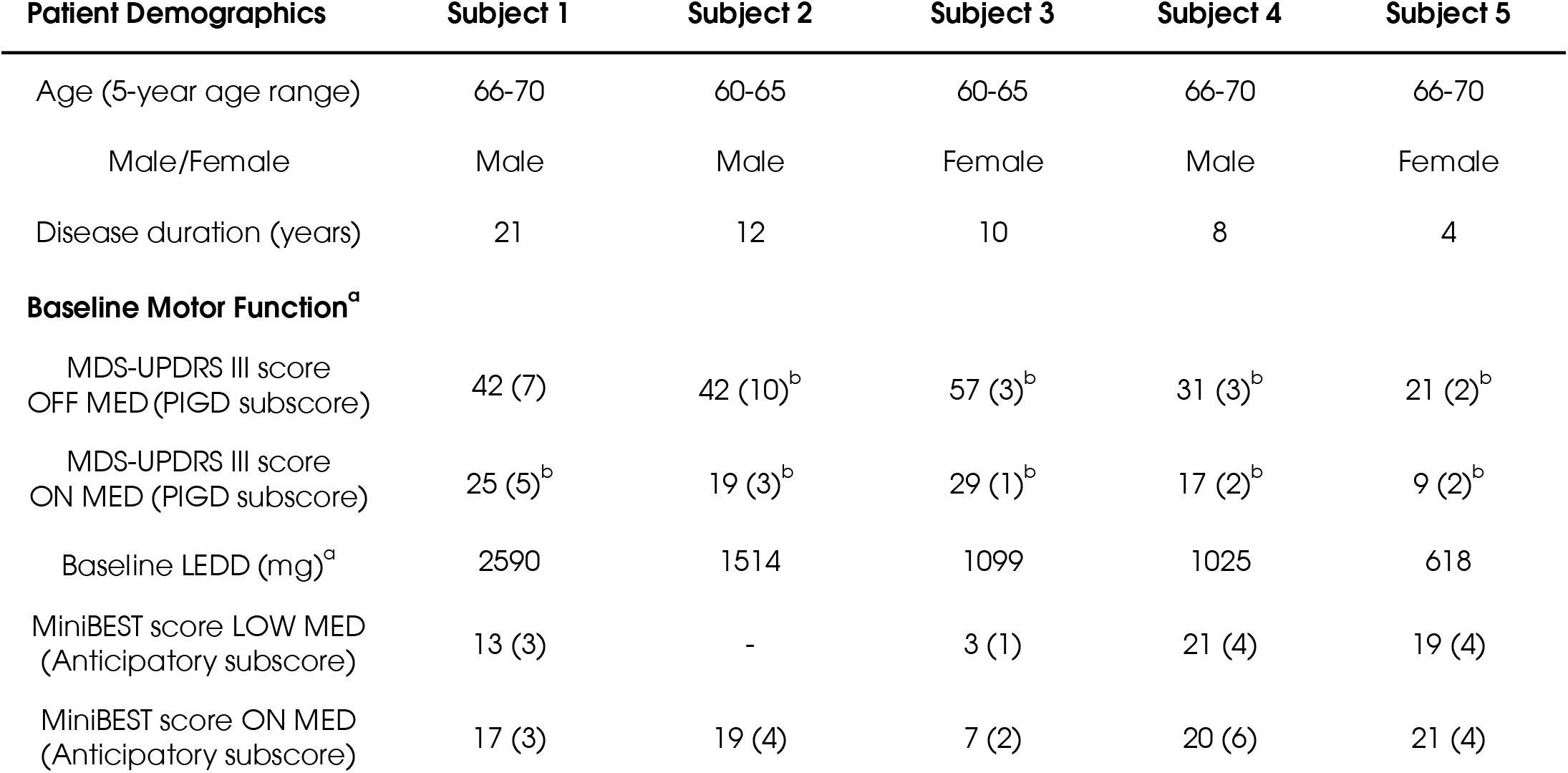
Subject demographics and baseline motor function *Legend*: ^a^Values taken from closest visit prior to DBS implantation; ^b^Test performed virtually by neurologist so rigidity and pull-test not examined. Abbreviations: MDS-UPDRS-III = Movement Disorders Society’s Unified Parkinson’s Disease Rating Scale, Part III - motor domain; PIGD = Posture Instability Gait Disorder (subscores from items 3.9: arising from chair, 3.10: gait, 3.11: freezing, 3.12: postural stability and 3.13: posture); LEDD = L-dopa equivalent daily dose; MiniBEST = Mini Balance Evaluation Systems Test.

#### Surgery

All subjects underwent implantation of quadripolar DBS leads targeting the pallidum (model 3387, Medtronic Inc.) and subdural cortical paddle electrodes (model 0913025, Medtronic Inc.) overlying the PMC and M1. Electrodes were connected to an investigational bidirectional neural stimulation device allowing chronic sensing of local field potentials (LFPs) in freely moving subjects (Summit RC+S, model B35300R, Medtronic Inc.).^20^ Blinding and randomization were not performed.

#### Electrode localization

Electrode localization was performed using established pipelines for depth and cortical electrodes by fusing preoperative All electrode locations were normalized into Montreal Neurological Institute space and visualized either on the FreeSurfer average cortical surface or a standardized subcortical atlas (Figure 1A).

**Figure 1.**
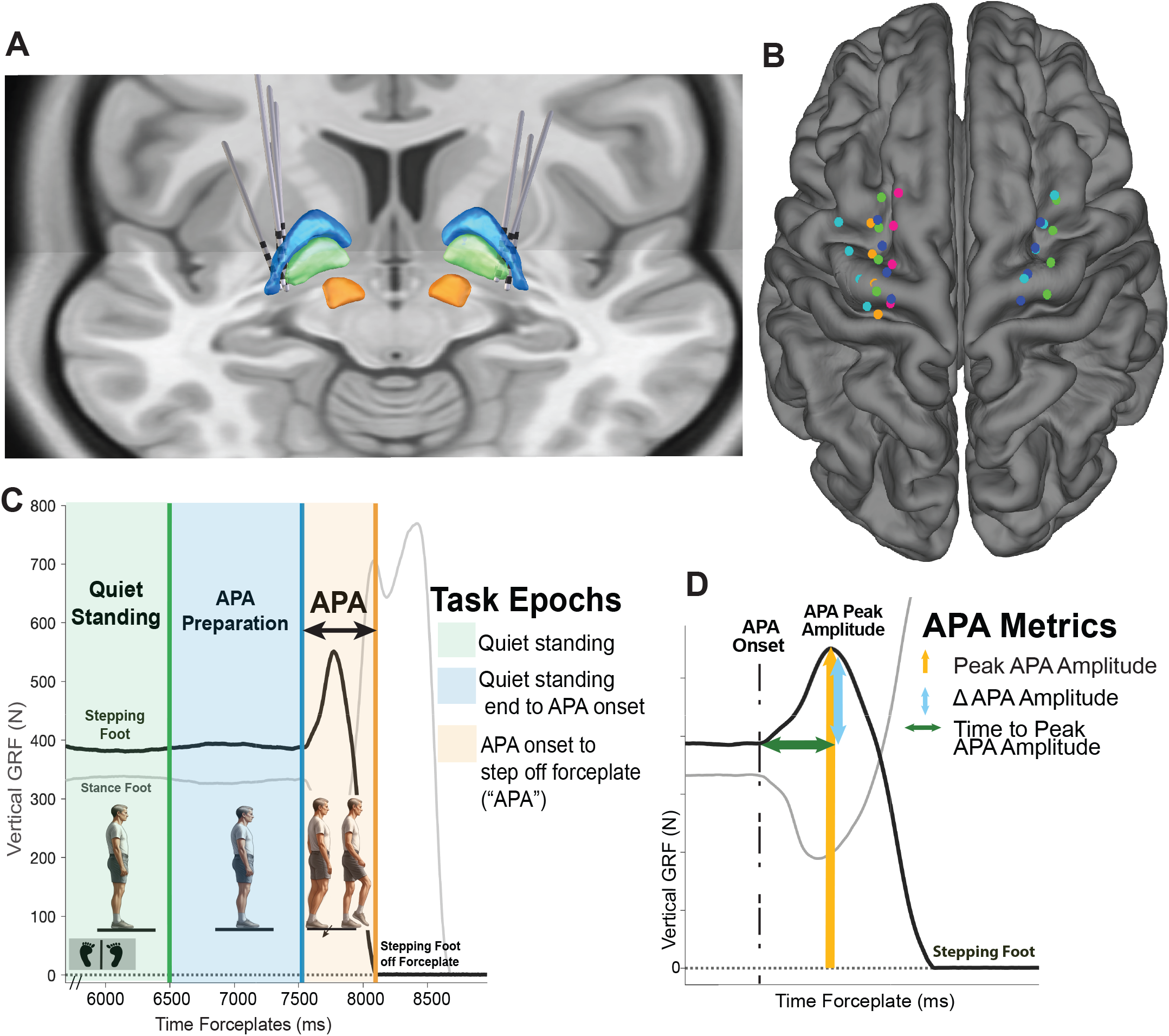
Recording electrode localization and gait initiation task scheme *Legend:* 1a: Aggregated subject DBS leads targeting the globus pallidus internus (green) and globus pallidus externus (blue) and ECoG contacts targeting the premotor cortex and primary motor cortex . 1b: Task epochs marked for each gait initiation trial featuring varying postural control strategies: “Quiet standing,” “Quiet standing end to APA Onset” (APA preparation), and “APA Onset to Stepping Foot off the Forceplate” (APA). 1c: The task’s APA metrics assessed by the study both “ON” and “LOW” medication included peak and change in APA amplitudes, and time to peak APA amplitude. Abbreviations: GRF = ground reaction force; ms = milliseconds; APA = anticipatory postural adjustment.

### Clinical motor symptom outcome measures

Subjects received baseline gait and balance assessments 1-2 months prior to DBS surgery, including the Movement Disorders Society’s Unified Parkinson’s Disease Rating Scale (MDS-UPDRS) motor subscore assessed by a movement disorders neurologist, as well as the Mini Balance Evaluation Systems Test (Mini-BEST) by a physical therapist (Table 1). These tests were administered ON and LOW medication and OFF DBS; the “ON” medication state was approximately 30 minutes after the ingestion of subjects’ typical dose of Parkinsonian medication and confirmation of “ON” motor functioning. The “LOW” medication state was defined as the withholding of one or more medication doses, spanning from overnight to multiple hours across subjects depending on symptom tolerance and dosing regimen.

### Gait initiation task

Subjects performed gait initiation trials on twin AMTI force plates (1000 Hz sampling rates) using a self-selected stepping foot following a visual cue. Each trial began with the patient choosing their weight distribution with a foot on each force plate. Subjects were instructed to begin walking upon display of a green screen; no other feedback was provided to maximize naturalistic performance. Gait initiation trials were performed in 5-10 trial sessions depending on patient tolerance, with seated breaks between sets and as needed.

### Biomechanical data collection and processing

Ground reaction force data from the force plates’ z-axes (GRFz) were processed using a low-pass, 4^th^-order Butterworth filter with 50 Hz cutoff frequency.^21,22^ Kinematic data were also collected using wireless surface EMG (Trigno, Delsys Inc., Natik, MA) and an inertial measurement unit (IMU) system (Xsens, Movella, Netherlands). The IMU system consisted of 15 units placed in a standard configuration^23^ over the subjects for data synchronization with an automatic force plate trigger. EMG sensors were placed on top of subjects’ RC+S implants for LFP synchronization.

### Task data analysis

Force plate data were separated into epochs during the task using custom MATLAB scripts to demarcate a period of quiet standing, APA preparation, APA, and stepping (Figure 1B). These points were chosen to gain insight into the differing postural control requirements across the task, including static (quiet standing) and anticipatory postural demands and motor execution (APA and stepping).^24^

#### Quiet standing

Quiet standing start was defined as the last GRFz crossing of the subjects’ feet on the force plates prior to the “Quiet Standing End” mark; subjects were to stand as quietly as possible on the force plates. The “Quiet Standing End” mark was 1.5s before the subjects’ stepping foot completely left the force plate. (This “rule” was established as patients took 0.5-0.75s to perform the APA and step, creating a 0.75-1s *preparatory APA* period.) Trials were required to have a minimum of 3s of quiet standing (with 2.5s used for one subject due to difficulty with quiet standing).

#### APA preparation

Used as a proxy for APA planning, the period from “Quiet Standing End to APA Onset” included the cue (length and reasoning above). APA onset was marked as the instance when the stepping foot’s GRFz slope exceeded 3.5 times the standard deviation of its slope averaged during quiet standing (adapted from published methods; verified and adjusted by visual inspection such as when subjects did not exceed threshold).^3,25^

#### APA

APA spanned from APA onset to when the stepping foot completely left the force plate (GRFz crossed 0 N). If GRFz did not fully cross 0 N due to calibration, the local minimum closest to 0 N was used.

### APA metrics

APA metrics used to assess task quality and performance included: 1) Peak APA amplitude, normalized to participant bodyweight and used as a proxy for APA vigor; 2) Δ APA amplitude (the difference between peak and APA amplitude at onset), also normalized to bodyweight and used as a proxy for APA scaling; and 3) Time to peak APA amplitude, defined as the time between APA onset and APA peak amplitude (Figure 1C).

### Neural data collection and processing

All neural and biomechanical data were collected after the **subjects had been implanted with the RC+S device, prior to DBS being turned ON**. The interval between DBS surgery and data collection varied among subjects, from 14-44 days.

LFPs were recorded in a sandwich configuration from the following electrode pairs: +2-0 (more ventral, targeting the GPi) and +3-1 (more dorsal, targeting the GPe and striatum). ECoG potentials were recorded using contact pairs +9-8 (over M1) and +11-10, over PMC. LFPs were sampled at 500 Hz and initially preprocessed using a preamplifier high-pass filter of 0.85 Hz and a two-staged low-pass filter at 1700 Hz and 450 Hz. Accelerometry data from the RC+S device was sampled at 64 Hz. All data were extracted using open-source code (https://github.com/openmind-consortium/Analysis-rcs-data).^26^

Trial-by-trial neural data were aligned with biomechanics data using peaks in accelerometer data from the Xsens and Delsys systems and RC+S device. Alignments were verified visually, with a 3-sample difference allowed between accelerometry peaks in the alignments. Dropped neural data packets in the RC+S recordings were also identified (0.01s threshold); no trials were removed due to dropped packets.

### Data analysis

All task epochs for each trial were visually inspected for correction. All gait initiation trials analyzed featured neural data from the contralateral hemisphere to the stepping leg.. Trials were excluded if patient was unable to perform quiet standing due to dyskinesia, lacked APA, or had multiple APAs without quiet standing. . For Subjects #1, 2, 4, and 5, 89% (74/83) of trials met the criteria and were analyzed; 37% (24/65) of trials were analyzed for Subject #3 due to difficulties performing the task.

*Artifact rejection:* Data were processed using a multitaper spectral transform (16 voices per octave, 75-150 Hz gamma filter limits, 60-time bandwidth),^27,28^ then normalized using median-based z-scoring. Data were blanked over time intervals where gamma power exceeded a set threshold of deviations. Thresholds and potential artifacts were evaluated for appropriateness with visual consideration regarding the overall data “noise,” waveform of potential artifacts, and regularity. Subject #4 had three artifacts that were removed. Subject #3’s data featured continuous artifact-appearing bands across all time points. These bands were present at the GPi (68 Hz, 131 Hz, and 200 Hz), and M1 (75 Hz) while ON medication, and at GPi (66 Hz, 70 Hz, 131 Hz, 200 Hz), and M1 (74 Hz) while LOW medication. Data were blanked at the frequencies above and +/- 2 additional frequencies for this subject’s trials in group analyses.

*Signal processing*: Neural data for each trial were analyzed using two built-in MATLAB signal processing functions for magnitude-squared coherence (“mscohere” function: 32 segment hamming window, 256 nfft, 90% overlap) and short-time Fourier transform (“spectrogram” function: 1s window, 90% window overlap, transform length 512 data points). Subject #4’s trials with artifacts were not included in coherence analyses for the implicated contacts due to their scaling. Data were filtered through a high pass, 4^th^-order Butterworth filter with 2 Hz cutoff to remove low frequency noise. Absolute epoch neural power and coherence were calculated over time and averaged across the canonical frequencies, with data normalized to the “Quiet Standing” epoch when inputted into the linear mixed model analyses. The canonical frequencies included theta (4-8 Hz), alpha (8-12 Hz), low beta (13-20 Hz), high beta (20-30 Hz), low gamma (30-50 Hz), and broadband gamma (50-200 Hz). Epoch power results were visualized and reflect the median power spectral density (PSDs) and median absolute deviations using MATLAB function “pwelch” (segment length 128 samples and 90% overlap). Subject #3’s data were notch filtered at the frequencies above for PSDs.

### Statistical Analysis

APA metrics were compared between LOW and ON medication states using two-tailed Wilcoxon rank sum tests in R (Table 2). Data were evaluated for normality prior to Wilcoxon rank sum testing using the Shapiro-Wilk test. Neural data were compared between three gait initiation task epochs (quiet standing, APA preparation, and APA) using Kruskal-Wallis (two-tailed) and post-hoc Dunn’s tests, with multiple corrections via the Benjamini-Hochberg method. LMMs were constructed using combined, normalized data (percent change) for all subjects in R using the “lme4” package,^29^ with backwards model selection using the “step” function and APA metrics as outcomes, All neural data and medication predictors for a single task epoch were ran at a time, allowing for comparisons between model performance and predictor significance between epochs. Fixed predictors inputted into the LMM included neural data for all frequency bands at each recording contact under both medication states; subjects were inputted as random effects. Due to overall trial number and increasing model complexity with greater random effects, only subjects were included as random effects. All final models reached suitable convergence.

**Table 2:** Gait initiation trials summary data *Legend:* ^a^Metric normalized to subject body weight in kilograms (kg); **\* “LOW” vs “ON” medication APA metrics significantly different (p < 0.05).** Abbreviations: GI = gait initiation; APA = anticipatory postural adjustment; N = Newtons; ms = milliseconds; std = standard deviation.

| Gait Initiation Data | Subject 1 | Subject 2 | Subject 3 | Subject 4 | Subject 5 |
| --- | --- | --- | --- | --- | --- |
| Stepping foot for all GI trials | Right | Right | Left | Left | Right |
| Neural hemisphere analyzed | Left | Left | Right | Right | Left |
| Number of trials analyzed<br>ON MED/LOW MED | 4/3 | 5/6 | 13/11 | 13/14 | 14/15 |
| Average peak APA<br>amplitude (std)<br>LOW MED/ON MED<sup>a</sup> (N/kg) | 6.45 (3.0)/<br>6.96 (0.9) | **6.86 (0.2)/**<br>**7.96 (0.5)<sup>\*</sup>** | 6.13 (1.6)/<br>5.43 (1.8) | 5.10 (0.9)/<br>6.14 (1.5) | 6.77 (0.4)/<br>6.82 (0.4) |
| Average $\Delta$ APA amplitude<br>(std)<br>LOW MED/ON MED<sup>a</sup> (N/kg) | 2.53 (2.3)/<br>2.04 (0.7) | **1.74 (0.3)/**<br>**2.23 (0.3)<sup>\*</sup>** | 2.18 (0.9)/<br>2.18 (1.0) | **0.73 (0.4)/**<br>**1.39 (0.8)<sup>\*</sup>** | 1.49 (0.4)/<br>1.40 (0.3) |
| Average time to peak APA<br>(std)<br>LOW MED/ON MED (ms) | 229 (48)/<br>318 (174) | **227 (34)/**<br>**277 (21)<sup>\*</sup>** | 240 (74)/<br>279 (95) | 283 (106)/<br>247 (84) | 275 (56)/<br>269 (47) |

LMMs were assessed for quality using Akaike Information Criterion (AIC), with Q-Q plots for residuals. All fixed effects in the final models were evaluated for collinearity using variance inflation factor (VIF). Model results in the main text include all significant predictors from backwards selection for each APA metric from the “APA” epoch, with additional epoch model summaries and model data in the supplemental figures (Figure 6; Supplemental Fig. 8A-C).

## Results

### Levodopa has variable effect on APAs during gait initiation

Five subjects (3 males and 2 females) with PD participated in the study (Table 1). Ninety-eight gait initiation trials were included for analysis which met criteria (Table 2). Neural data were analyzed from the brain hemisphere contralateral to the stepping foot, which was the left hemisphere in Subjects #1, #2, and #5 and right hemisphere in Subjects #3 and #4. Trials performed in the LOW and ON medication states were relatively matched within subjects and two subjects (#2 and #4) demonstrated significantly decreased APA amplitudes in the LOW state. Average APA peak amplitude Newton (N)/ kilogram (kg) across subjects ± standard deviation was 6.139 ± 1.306 N/kg while LOW medication and 6.397 ± 1.445 N/kg while ON (p = 0.19). Average Δ APA amplitude across patients was 1.524 ± 0.925 N/kg in the LOW medication state and 1.744 ± 0.790 N/kg while ON (p = 0.12). Average time to peak APA across patients was 261 ± 76 ms in the LOW state and 270 ± 83 ms while ON (p = 0.46).

### Gait initiation is associated with decreased beta and increased gamma power in the pallidum

To evaluate signals from the pallidum during gait initiation, we analyzed the absolute mean spectral power across the following epochs of the gait initiation task: quiet standing, APA preparation, and APA, using grouped subject data and Kruskal-Wallis and Dunn’s post-hoc testing with Benjamini-Hochberg corrections (Figures 3-4).

**Figure 2:**
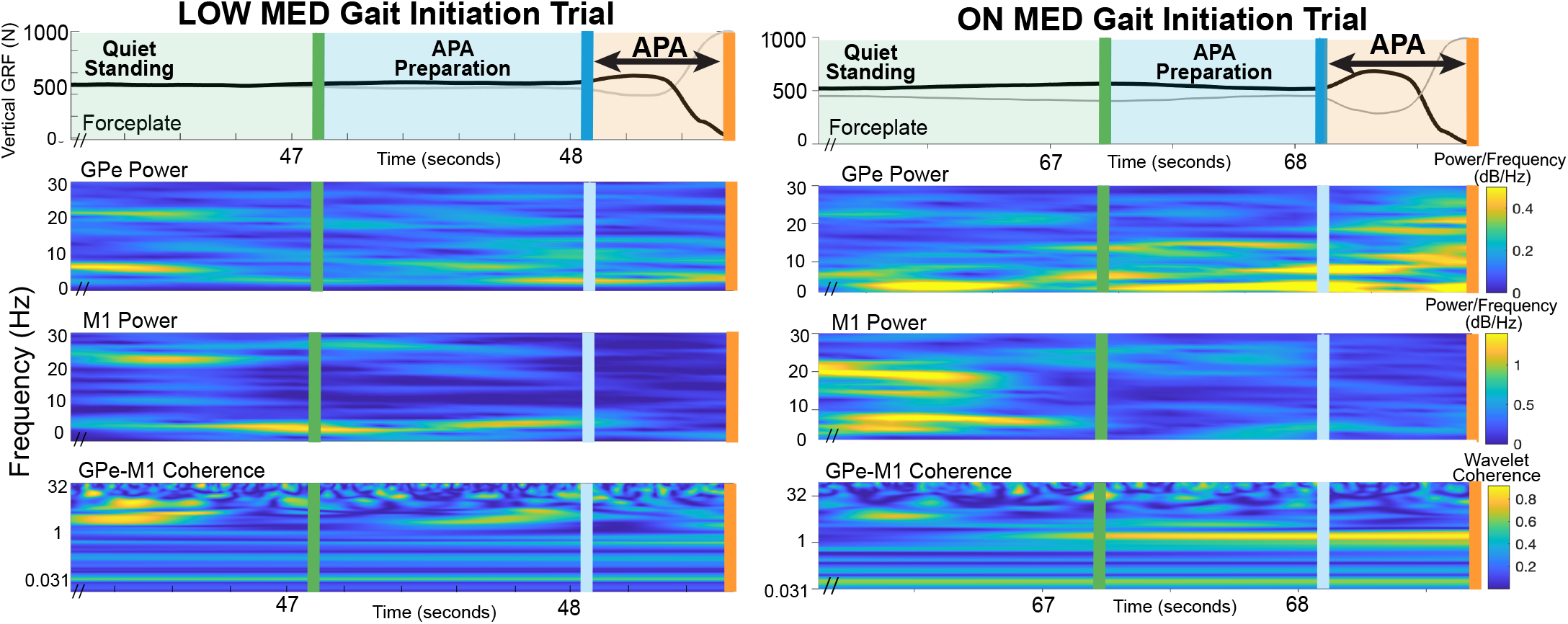
Sample pallidal and cortical spectrogram and coherogram for a gait initiation trial “LOW” and “ON” medication states *Legend:* Sample subject’s “LOW” (left panel) vs “ON” (right panel) single-trial gait initiation data reflecting task epochs and biomechanical data (top plot), absolute neural power (middle plot) via spectrogram, and absolute wavelet coherence (bottom plot). Neural power and coherence were averaged within each epoch for all trials and contacts.

**Figure 3:**
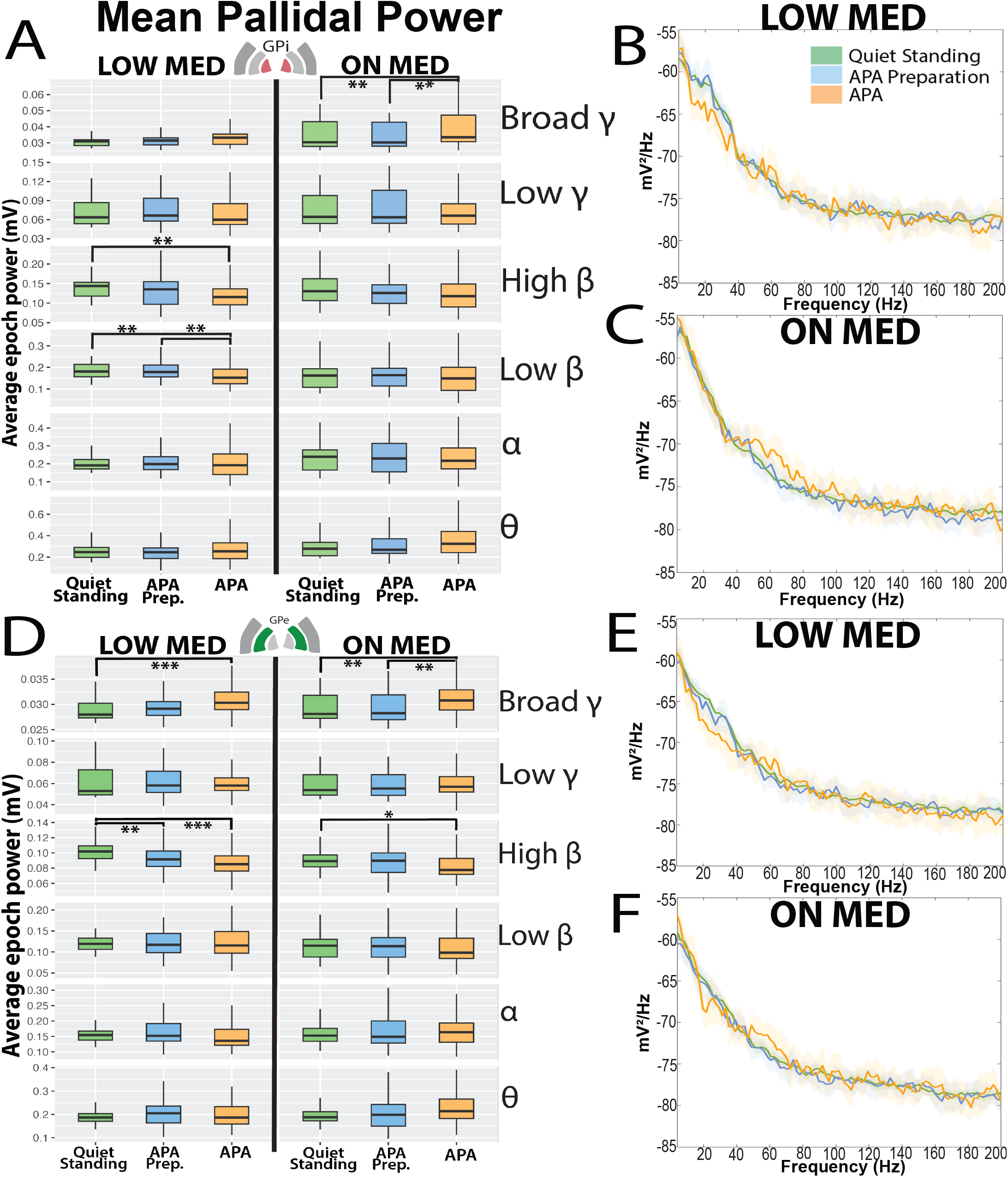
Pallidal power changes across gait initiation in LOW and ON medication states *Legend:* Grouped subject data at GPi (panels A-C) and GPe (panels D-F): B/C and E/F reflect median power spectra from all subjects in the “LOW” and “ON” medication states during the three primary task epochs: Quiet standing (green), APA preparation (blue) and APA (orange). A and D: Boxplots demonstrate grouped average power from all subjects from the “spectrogram” function during each epoch across the canonical frequencies. Significance between epochs is denoted with * p< 0.05, ** p<0.01, ***p<0.001 following Kruskal-Wallis and Dunns testing with multiple corrections. Boxplots created with upper whisker representing 1.5 * interquartile ratio (IQR) past the 3^rd^ quartile and lower whisker representing 1.5 * the IQR below the 1^st^ quartile. Significance is seen among power modulation at β and γ frequencies depending on contact location under both medication states. Abbreviations: GPi = globus pallidus internus, GPe = globus pallidus externus, Hz = Hertz; mV = millivolts; APA = anticipatory postural adjustment; θ = theta, α = alpha, β = beta, γ = gamma.

**Figure 4:**
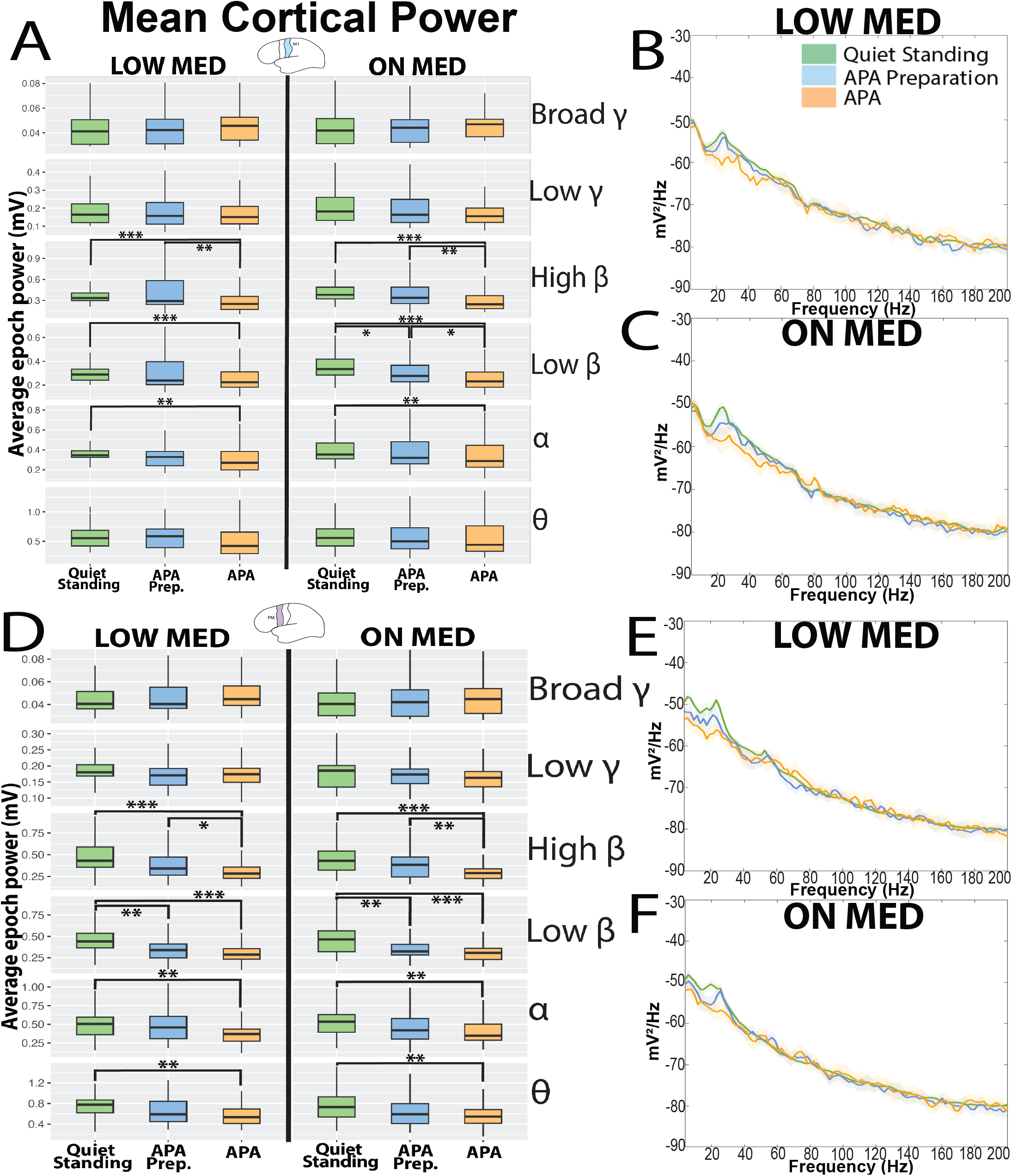
Motor cortex power changes across gait initiation epochs in LOW and ON medication states *Legend:* Grouped subject data at M1 (panels A-C) and PMC (panels D-F): B/C and E/F reflect median power spectra from all subjects in the “LOW” and “ON” medication states during the three primary task epochs: Quiet standing (green), APA preparation (blue) and APA (orange). A and D: Boxplots demonstrate grouped average power from all subjects from the “spectrogram” function during each epoch across the canonical frequencies. Significance between epochs is denoted with * p< 0.05, ** p<0.01, ***p<0.001 following Kruskal-Wallis and Dunns testing with multiple corrections. Boxplots created with upper whisker representing 1.5 * interquartile ratio (IQR) past the 3^rd^ quartile and lower whisker representing 1.5 * the IQR below the 1^st^ quartile. Significance is seen among power modulation at θ, α, and β frequencies depending on contact location under both medication states. Abbreviations: M1 = primary motor cortex, PMC = premotor cortex, Hz = Hertz; mV = millivolts; APA = anticipatory postural adjustment; θ = theta, α = alpha, β = beta, γ = gamma.

While LOW medication, power changed across the task in the pallidum for beta (β) and broadband gamma (γ) frequencies. In the GPi, median low β power showed significant progressive decreases from quiet standing (median [interquartile range]: 0.181 [0.152-0.210] mV) to APA preparation (0.178 [0.150-0.206] mV) to APA (0.152 [0.118-0.186] mV; standing vs APA, and APA preparation vs APA, p<0.01; Figure 3). GPi high β power significantly decreased between quiet standing (0.144 [0.127-0.162] mV) and APA (0.115 [0.095-0.136] mV; standing vs APA, p<0.01, Figure 3). High β power in the GPe also demonstrated significant decreases from quiet standing (0.102 [0.094-0.111] mV) to APA preparation (0.092 [0.082-0.102] mV), and between quiet standing and APA (0.085 [0.075- 0.095] mV; standing vs APA preparation p<0.01, standing vs APA p<0.0001, Figure 3). Interestingly, GPe broadband γ power significantly increased from standing (0.028 [0.027- 0.030] mV) to APA (0.030 [0.029-0.032] mV, p<0.001, Figure 3).

While ON medication, the pallidum exhibited slightly different modulation. Mainly, GPi no longer showed significant β power decrease across the task, presumably due to medication’s effects on decreasing resting β synchrony (median low β power: standing: 0.162 [0.119-0.205], APA preparation: 0.164 [0.123-0.205], and APA: 0.149 [0.096-0.202] mV; p = 0.5-1.0; median high β power: standing: 0.131 [0.103-0.159], APA preparation 0.126 [0.102-0.150], APA 0.118 [0.089-0.148] mV, p = 0.1-0.2, Figure 3). The GPi also demonstrated broadband γ power increase during APA (0.033 [0.025-0.042]) compared to APA preparation and standing (0.030 [0.022-0.038] mV, p<0.01, Figure 3). Levodopa did not significantly alter the oscillatory dynamic of the GPe across the task, as it showed similar high β power decrease during APA (0.0.78 [0.068-0.089]) compared to quiet standing (0.089 [0.081-0.097] mV, p<0.05, Figure 3). as well as similar broadband γ power increase with APA (0.031 [0.029-0.033] mV) compared to standing and APA preparation (0.028 [0.026- 0.031] mV, p<0.01, Figure 3). In summary, APA during gait initiation is associated with overall pallidal β power decrease and broadband γ power increase, with levodopa having more influence on these changes in the GPi than GPe.

### Beta band power in motor and premotor areas may play different roles in APA

At the cortex, task-related theta (θ), alpha (α), and β power decreased from quiet standing to APA irrespective of medication state. In the LOW medication state, M1 α and low β power significantly decreased from quiet standing to APA (α power: standing: 0.345 [0.312-0.379] mV, APA: 0.269 [0.177-0.362] mV, p<0.01, Figure 4; low β power standing: 0.288 [0.242-0.335] mV, APA: 0.223 [0.158-0.288] mV, p<0.001, Figure 4). Interestingly, M1 high β power decreased from quiet standing (0.334 [0.280-0.389] mV) to APA (0.251 [0.159-0.343] mV (p<0.0001), but also showed a significant decrease from APA preparation (0.292 [0.122-0.463] mV) to APA (p<0.01, Figure 4), indicating its role in APA execution. The ON medication state produced similar trends of α and β amplitude decrease during APA compared to quiet standing (Figure 4).

We found greater task-related power modulation during APA preparation in the PMC compared to M1. In the LOW medication state, PMC θ and α power significantly decreased between quiet standing to APA (θ power standing: 0.775 [0.650-0.901] mV, APA: 0.535 [0.394-0.676] mV, p<0.01; α power standing: 0.504 [0.387-0.622] mV, APA: 0.369 [0.289-0.450] mV, p<0.01; Figure 4). Strikingly, low β and high β power dynamics in the PMC appeared to differentiate between APA preparation and execution. Low β power in the PMC significantly decreased from quiet standing (0.441 [0.356-0.526] mV) to APA preparation (0.339 [0.259-0.420] mV, p<0.01) and quiet standing to APA (0.286 [0.225-0.348] mV, p<0.0001), but not from APA preparation to APA, suggesting that low β change may signify a preparation signal. PMC High β power exhibited similar decreased between quiet standing to APA and APA preparation to APA (Quiet standing: 0.431 [0.315-0.548] mV, APA preparation: 0.341 [0.237-0.445] mV, APA: 0.284 [0.220-0.349] mV, standing vs APA, p<0.001; APA preparation vs APA, p<0.05; Figure 4). Medication did not alter these observations for the PMC, as the ON medication state displayed the same significant trends across the task.

Despite individual power modulation variation across the task, all subjects showed the similar trend of pallidal β band power decrease and γ band increase during APA compared to quiet standing (Supplemental Figs. S1-5). Cortical responses were more variable, though most subjects showed lower-frequency (θ, α, β) power decrease with broadband γ power increase while LOW medication (Supplemental Figs. 1-5). The two individuals whose APAs improved with medication (Subjects #2 and #4) showed no marked difference in their gait- initiation related power modulation in the ON state compared to other subjects.

**Figure 5:**
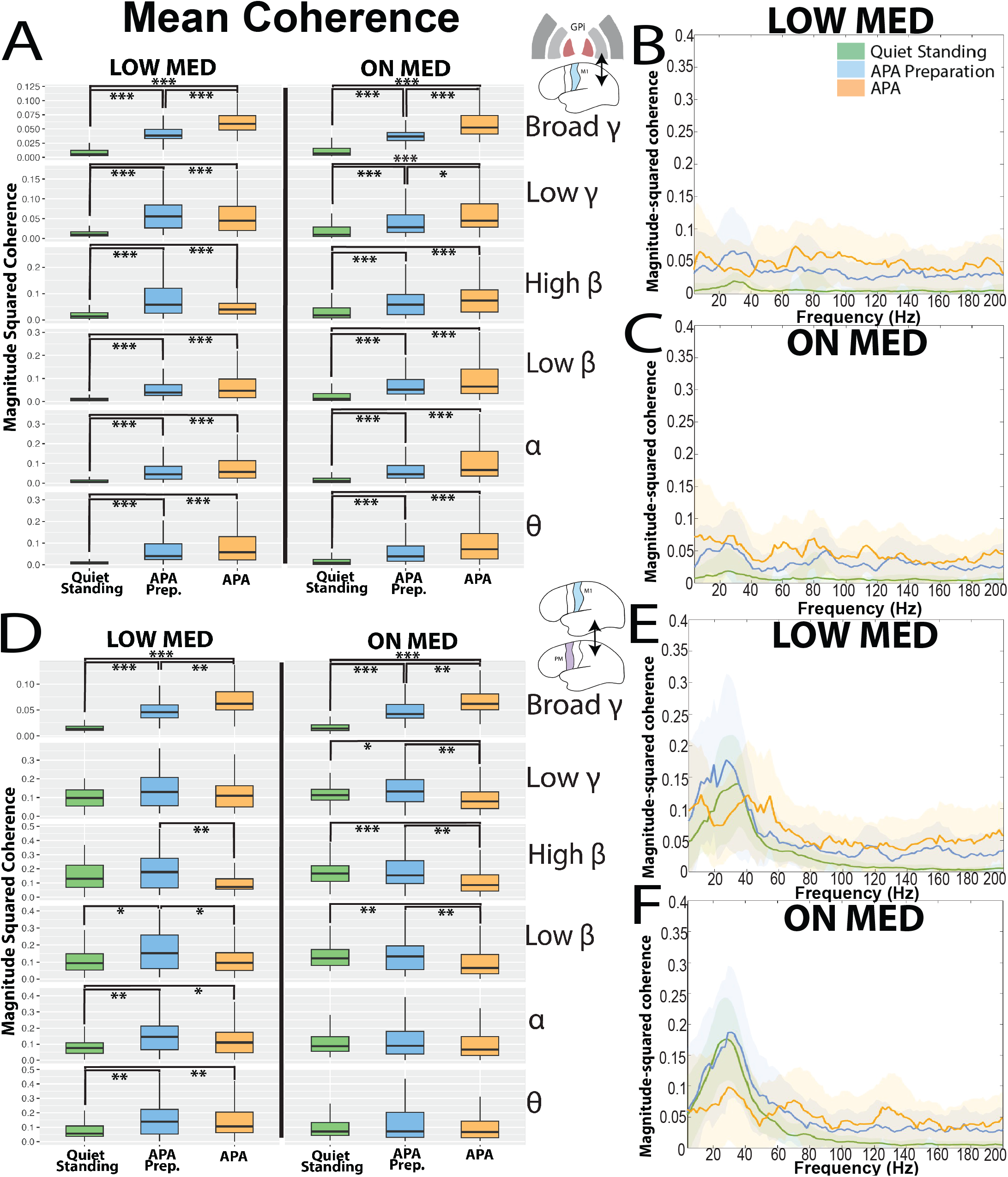
Pallidal-cortical coherence changes during gait initiation epochs in LOW and ON medication states *Legend:* Grouped subject data across GPi-M1 (panels A-C) and M1-PMC (panels D-F): B/C and E/F reflect median magnitude-squared coherence and median absolute deviations from all subjects in the “LOW” and “ON” medication states during the three primary task epochs: Quiet standing (green), APA preparation (blue) and APA (orange). A and D: Boxplots represent average magnitude-squared coherence during each epoch across the canonical frequencies. Significance between epochs is denoted with * p< 0.05, ** p<0.01, ***p<0.001 following Kruskal-Wallis testing and multiple corrections. Boxplots created with upper whisker representing 1.5 * the interquartile ratio (IQR) past the 3^rd^ quartile and the lower whisker representing 1.5 * IQR below the 1^st^ quartile. Significant task-related coherence modulation is demonstrated under both medication states and frequencies. Abbreviations: GPi = globus pallidus internus, M1 = primary motor cortex; PMC = premotor cortex, APA = anticipatory postural adjustment; θ = theta, α = alpha, β = beta, γ = gamma.

### Gait initiation involves increased pallidocortical coherence but decreased beta premotor-motor cortex coherence

To evaluate network changes between pallidal and cortical areas during gait initiation, we analyzed the absolute magnitude-squared coherence from all subjects across the task, using Kruskal-Wallis and Dunn’s post-hoc testing with Benjamini-Hochberg corrections.

Widespread coherence increase was seen across the task between pallidal-cortical regions, regardless of medication state. For GPi-M1, GPi-PMC, GPe-M1, and GPe-PMC, coherence significantly increased between quiet standing and APA preparation, and quiet standing compared to APA, under both medication states and across all canonical frequency bands. As a representative example, GPi-M1 low β coherence increased from quiet standing (LOW medication: 0.009 [0.004-0.014], ON: 0.011 [0-0.026]) to APA preparation (LOW medication: 0.039 [0.016-0.062], ON: 0.051 [0.019-0.083]; p<0.0001), and especially increased between quiet standing and APA (LOW medication: 0.047 [0.007-0.088], ON: 0.065 [0.01-0.12] p<0.0001; Figure 5, Supplemental Figures 6-7).

Interestingly, PMC-M1 β coherence exhibited more dynamic changes compared to that of pallidocortical during gait imitation. As an example of this in the LOW medication state, median low and high β coherence first increased during APA preparation and then decreased with APA (low β coherence standing: 0.093 [0.046-0.141], APA preparation:

0.151 [0.053-0.249], APA: 0.095 [0.045-0.145], standing vs. APA preparation and APA preparation vs APA, p<0.05; high β coherence standing: 0.131 [0.053-0.209], APA preparation: 0.177 [0.072-0.282], APA: 0.072 [0.035-0.110]; APA preparation vs APA p<0.01; Figure 5). These trends were similar in the ON medication state (For example, median ON high β coherence during standing: 0.167 [0.113-0.222], APA preparation: 0.154 [0.074-0.235], and APA: 0.085 [0.027-0.144]; standing vs APA p<0.001, APA preparation vs APA p<0.01, Figure 5).

To summarize, there were widespread increases in coherence between pallidal-cortical regions under both medication states across all canonical frequency bands, but intracortical β coherence decreased with APA execution.

### APA scaling is influenced by pallidal gamma power, while APA timing is driven by pallidocortical coherence

To identify oscillatory features which are most predictive of APA amplitude, scaling, and timing, we built separate linear mixed models (LMMs) using normalized neural power and magnitude-squared coherence recorded from the APA epoch (data normalized to the quiet standing epoch for each trial) (Figure 6, Supplemental Table 1).

**Figure 6:**
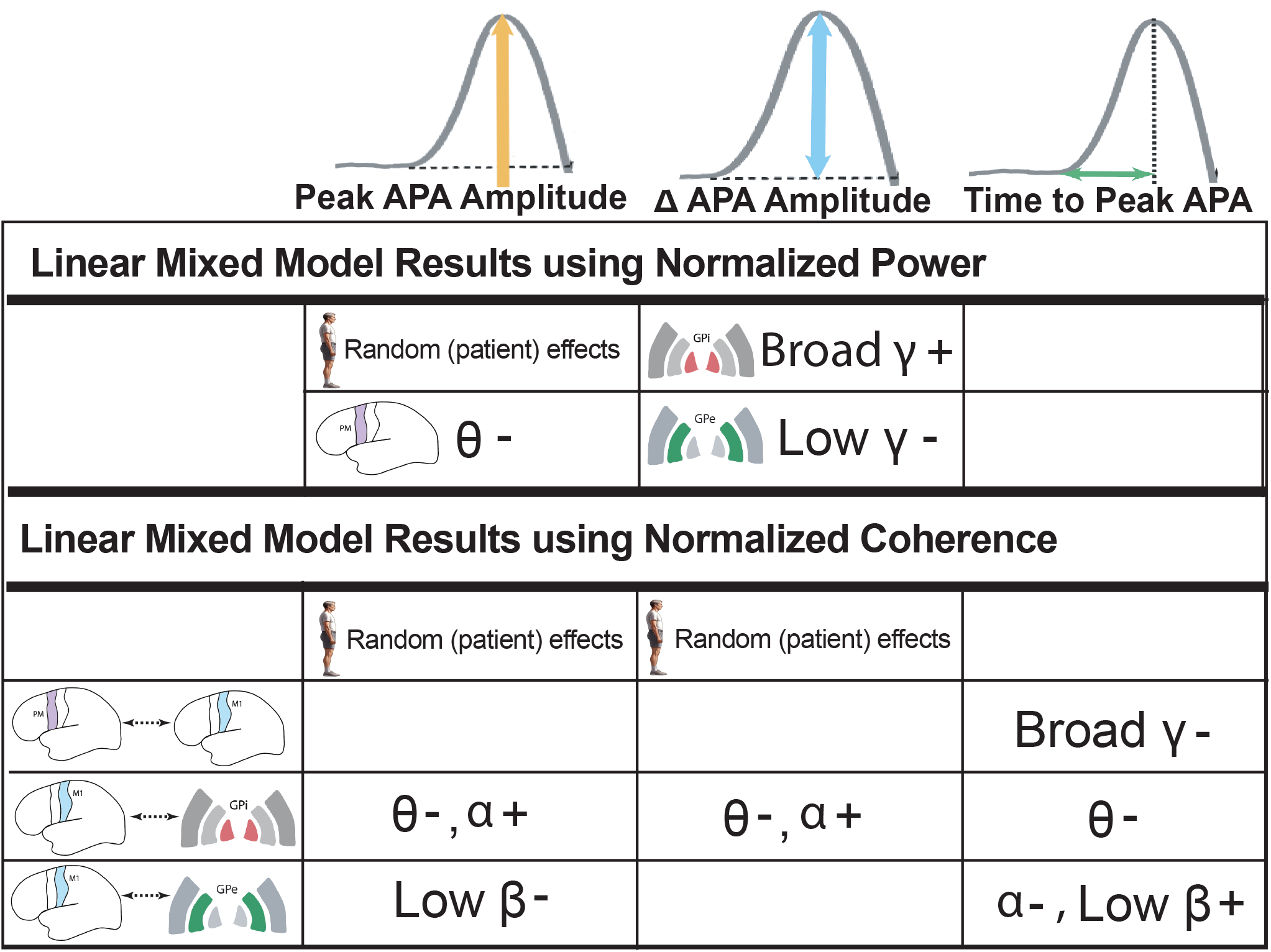
Optimized linear mixed model results for APA metrics from “APA” epoch. *Legend:* Linear mixed model results using normalized epoch power and coherence to predict APA metrics of amplitude and timing. The table provides significant model predictors and their relationship to each APA outcome following backwards model selection from the “APA” epoch. **+** : positive relationship between APA outcome and neural data; **-** : negative (inverse) relationship. Note: GPi-GPe coherence predictors were not included due to the likelihood of signal bias due to the sandwich recording configuration. Abbreviations: APA = anticipatory postural adjustments; GPi = globus pallidus internus, GPe = globus pallidus externus, M1 = primary motor cortex, PMC = premotor cortex; θ = theta, α = alpha, β = beta, γ = gamma.

PMC θ power (estimate = -0.011, t = -2.38, p<0.05) was negatively correlated with peak APA amplitude and the random effects of subject variability were a significant predictor of this metric. For Δ APA, GPi broadband γ power (estimate = 0.018, t = 2.47, p<0.05) was found to be positively correlated, while GPe low γ power (estimate = -0.014 t = -2.64, p<0.01) negatively correlated with APA scaling. The time to peak APA amplitude metric did not include any significant power predictors following backwards model selection. Interestingly, levodopa did not bear any significant impact on these APA metrics following backwards model selection and optimization using inputs of neural power (Figure 6).

Using normalized coherence as inputs for the LMM, GPi-M1 θ (estimate = -0.00013, t = -2.77, p<0.01) and GPe-M1 low β coherence (estimate = -0.0000382, t = -2.36, p<0.05) were found to be inversely correlated, while GPi-M1 α coherence (estimate = 0.00023, t = 3.00, p<0.01) was positively correlated with peak APA amplitude. The random effects of subject variability were also significant in this model. Predictors of Δ APA amplitude in the optimized LMM included decreased GPi-M1 θ coherence (estimate = -0.000095, t = -3.50, p<0.001) and increased α coherence (estimate = 0.00013, t = 2.79, p<0.01). The random effects of subject variability were again significant in this model. Interestingly, time to peak APA amplitude appeared to be driven by several coherence measures without significant impact from subject variability. These included: intracortical broadband γ coherence (estimate = -0.000050, t = -2.15, p<-.05), as well as GPi-M1 θ coherence (estimate = - 0.0000029, t = -2.10, p<0.05), which were shown to be negatively correlated with time to peak amplitude. Other predictors included GPe-M1 α (estimate = -0.0000011, t = -3.28, p<0.01) and low β coherence (estimate = 0.0000028, t = 2.78, p<0.01), which negatively and positively, respectively, influenced the metric. Intrapallidal (GPi-GPe) coherence were not used when assembling LMMs due to the likelihood of artificially-elevated coherence measurements from overlapping signals recorded from DBS leads in a sandwich configuration.

In summary, despite individual variabilities which influence APA amplitude, APA scaling (Δ APA amplitude) is largely affected by pallidal γ power, whereas APA timing is driven by coherence across the PMC-M1 and M1-pallidal networks. Supplemental Table 1 includes all model statistics including additional models built from the APA preparation epoch.

## Discussion

To our knowledge, this is the first study that examines pallidal-cortical network interactions during a gait initiation task in humans. By characterizing the dynamics of pallidal and motor cortical field potential and coherence changes on timing and amplitude metrics of APAs during gait initiation, we gain insight in how these brain regions and their communication influence gait initiation performance and quality. First, we demonstrate that gait initiation in Parkinson’s disease involves decreased, incremental low frequency power and increased broadband γ power modulation across the task. Second, we characterized coherence dynamics across the task, which showed widespread, increased pallidal-cortical coherence across all canonical frequencies with task progression, except decreased intracortical β coherence with APA and stepping. Lastly, our results indicate that pallidocortical and intracortical coherence is highly influential in predicting APA timing, while pallidal γ power predicts APA scaling. Interestingly, levodopa medication seemed to have little effect on these results.

### Incremental beta and gamma power modulation during gait initiation is largely unaffected by levodopa

A main finding of this work characterizes cortical and pallidal power modulation across a human, bipedal postural transition in people demonstrating gait and balance issues. Our results reflect pallidal β power decreases from quiet standing, APA preparation, and APA with stepping. Broadband γ power similarly increased across the task. This modulation was reflected at the GPe regardless of levodopa state, with decreased modulation at the GPi while ON medication. At the motor cortex, decreased lower-frequency oscillations (θ, α, β) occurred with task progression regardless of levodopa state. While research in healthy adults has suggested bipedal gait initiation to be preplanned via cortico-thalamo-basal ganglia circuits, little information exists quantifying the magnitude and timing of the neural changes coinciding with these events in people with PD, allowing for greater understanding of sources of dysfunction.

Much evidence supports pallidal β desynchronization occurring with human movement.^30^ As the GPi is the primary output of the basal ganglia, β desynchronization there is thought to mediate motor program flexibility by enhancing communication with cortico- thalamic targets during motor program switching for task execution.^31^ Studies utilizing simple motor tasks such as upper-extremity movements have illuminated presumably physiologic features of pallidal β desynchronization, including its relation to motor processing and movement speed.^30,32,33^ Our results help to validate the existence of pallidal β desynchronization with gait initiation onset, and provide evidence of task-related, graded, β modulation during scaling of motor responses during postural transitions.

Prior research has suggested pallidal γ synchronization to be a “pro-kinetic” signal,^30,34^ however, limited evidence suggests how its modulation is affected with movement scaling. A previous study reported “stepwise” broadband γ modulation in people with dystonia which correlated with movement amplitude (larger movements demonstrated stronger synchronization).^32^ Another study in people with PD performing simple hand movements reported maximal STN broadband γ power to coincide with peak movement velocity, relative to movement onset and amplitude.^35^ Our results provide additional insight, suggesting pallidal β desynchronization and broadband γ synchronization during a weightbearing task may influence the force generated through increased motor unit recruitment and overall task demands between quiet standing and APA/stepping.

These trends were also reflected at the cortex. EEG studies associated central lower-frequency (8-30 Hz) desynchronizations with normal gait initiation in healthy adults.^14^ This modulation occurred 800ms before APA onset, with synchronization afterwards until 400ms before stepping.^14^ Intriguingly, our results demonstrate lower-frequency desynchronizations in people with PD relative to quiet standing at the cortex through *both* gait initiation preparation and execution. More specifically, we identify that low and high β oscillations may play different roles in the gait initiation task. Our data suggest that low β desynchronization in the PMC may represent a preparatory signal for the postural adjustments prior to high β desynchronization in both the PMC and M1 that is found during APA execution. We hypothesize that these prolonged beta desynchronizations found in the cortex of PD patients may represent a compensatory mechanism required to drive gait initiation—through suppression of excessive β synchronization throughout the pallidal-motor network.

Levodopa was shown in our results to only influence task-related power modulation at the GPi but not significantly at the GPe and motor cortical areas. As the major basal ganglia outflow, dopamine depletion in the Parkinsonian state has been shown to increase resting GPi β oscillations,^36–39^ and in the GPe.^40,41^ Interestingly, we found levodopa only decreased pallidal β power during quiet standing and increased pallidal γ gamma throughout all epochs of gait imitation. Levodopa did not have any significant effects on the motor cortex and the GPe, suggesting that it may have limited effects on pathological synchronization during this complex motor task, thereby providing a potential mechanism for why postural instability remains a difficult symptom to effectively treat in this population.

### Gait initiation features widespread, progressive increases in coherence across cortical-subcortical regions

Our group-level results provide novel characterization of cortico-pallidal and intracortical coherence during gait initiation. Coherence generally increased for θ, α, β, and γ frequencies as the task progressed from quiet standing to APA and stepping, regardless of medication state. Intra-cortical β coherence also generally increased from quiet standing to APA preparation under both medication states, before decreasing upon APA and stepping.

Cortico-pallidal coherence is thought to be a proxy for functional connectivity during movement and is often elevated in PD.^38^ β coherence is assumed to have a similar physiological role as β power for maintaining the motor program, with hypersynchrony thought to impair information flow within motor circuits. Our results suggest coherence to be low during quiet standing, then increase incrementally with movement scaling. We hypothesize that in our subjects, increased coherence may have accompanied cortical engagement of the palladium to release tonic inhibition and facilitate movement scaling during APA and stepping. It is also hypothesized that a breakdown in preplanning or loss of automaticity in people with PD would require adjustments in motor signaling (i.e. increased reliance on “top-down” motor control) for compensation during task execution.

Other research has reported basal ganglia-M1 coherence to decrease with movement in people with PD and dystonia.^42,43^ This was hypothesized to allow the release of motor cortical activity for movement via a “GPi-gating” mechanism created by the interruption of GPi-cortical connectivity.^43^ Since our subjects have motor issues, it is unknown whether this elevated coherence represents physiological or pathological neural modulation. Many of these studies utilize non-weight-bearing tasks (e.g. button pressing), offering unknown application to weight-bearing and/or complex movements.^42,43^ We also observed increased broadband γ coherence across the task, which likely represents a prokinetic signal.^44^

Our results for intra-cortical coherence are similar, except β coherence under both medication states generally increased from quiet standing to APA preparation and decreased upon APA and stepping. Previous research has reported elevated cortical coherence across α, β, and γ frequencies, which was linked to postural control issues such as FoG in PD.^45^ If this finding is physiological, it suggests that successful gait initiation requires a decrease in β coherence during APA and stepping, reflecting “top-down” control from the motor cortical regions. This may serve as a “brake release” for the activation of motor programs at the pallidum for gait initiation, especially for movement scaling.

### Lower-frequency pallidal-motor cortical coherence has significant influence on APA scaling and timing

The LMM results reflect the significance of pallidal gamma power on APA scaling and coherence (both pallidal-cortical and intra-cortical) in influencing APA timing.

Specifically, APA scaling was highly influenced by increased GPi broadband γ and decreased GPe low γ activity. This follows what is known about the essential roles of the basal ganglia in movement scaling.

We found intra-cortical broadband γ coherence to have the largest (negative) relationship with time to peak APA amplitude. (E.g., elevated coherence results in a shorter time to peak amplitude). This is consistent with the theorized role of broadband γ frequencies in the motor cortex, which likely represent synchronized spiking activity leading to a prokinetic signal. It is assumed that more efficient and coordinated movements would result from the shared firing of neurons between the motor planning and motor execution regions, reflected through faster APA execution.

Additionally, APA timing was also influenced by lower-frequency (θ and α) M1- pallidal coherence, with greater coherence resulting in a shorter time to peak APA. M1- pallidal β coherence was also positively correlated with time to APA (greater coherence results in longer time to peak APA). Our findings are consistent with the theorized roles that coherence across these frequency bands play in motor circuits. As discussed earlier, α coherence is thought to mediate a motor “resting state”, with basal ganglia-cortical α coherence decreasing with simple movements.^42,43^ Similarly, θ coherence is thought to represent attention and sensory integration during motor tasks.^46^ As cued gait initiation is presumably more complex, our results suggest that θ and α M1-pallidal coherence may mediate the inhibition of task-irrelevant information while facilitating the integration of task-relevant feedback (e.g. error processing). This could result in more efficient motor scaling, as we observed. Our finding of increased β coherence resulting in a longer time to peak APA amplitude is consistent with its theorized role as a motor “holding” signal, likely due to M1 holding the pallidum in a synchronized state. Once released from the M1 “hold,” the pallidal prokinetic gamma oscillations regulates the scaling of the postural responses.

As our results are in individuals with gait initiation difficulties, it is unclear whether the modulation observed in coherence and power across the task are reflective of physiological phenomenon or dysfunctional processes. These results reinforce the importance of circuit connectivity and neural modulation in mediating PD-related motor symptoms, however, further investigation is required to untangle under which conditions, regions, and in which patients coherence switches from being “elevated and pathologic to “beneficial and/or compensatory.” Regardless, these findings validate ideas that transient network changes are critical for the preparation and execution of this task.

#### Limitations

A primary limitation is the small sample size due to the invasive and investigational nature of the neural device, which could limit the generalizability of results. Another limitation is the varied number of gait initiation trials analyzed for each subject due to individuals’ motor function and symptoms or fatigue, as well as minor trial elimination due to stepping with the opposite foot, lacking APA, or the presence of multiple APAs. Every attempt was made to utilize all trials while maintaining consistency across patients and task markings. Other limitations include subjects not being tested in a complete OFF medication state (due to safety concerns or subjects’ inability to perform the task), and the unknown transferability from a laboratory to a naturalistic environment. It is also important to acknowledge the patient-specific factors which contribute to the results, including differences in LEDDs, baseline motor function, PD motor classification, time since diagnosis (impacting levodopa responsiveness), neurophysiological differences (e.g. lead location, impedances, anatomical variance), and/or anthropometrics. Continued research across a larger patient population with varying characteristics would further validate our findings.

## Conclusions

Gait initiation is a fundamental, bipedal motor task requiring APA for effective performance. We used a novel approach to examine the corticopallidal interactions that modulate this task under different medication states. Our results contribute to the assembly of a conceptual framework for understanding the neural changes during complex motor activities and their relationship to task performance. Future research building upon this framework can lead to the development of effective interventions for treating dysfunction for this motor task.

## Data Availability

Data from this study can be made available upon reasonable request, following patient confidentiality and disclosure standards.

## Data Availability

Data from this study can be made available upon reasonable request, following patient
confidentiality and disclosure standards.

## Acknowledgements

We appreciate the subjects for participating in this study. Many thanks to Dr. Martina Mancini and Dr. Fay Horak for their contributions to the study design and methods. We also thank Dr. Abel Torres-Espin for his invaluable input on the linear mixed models.

## Supplementary Material

Supplemental items 1-8.

