## Supplemental Materials for "Pallidal and motor cortical interactions determine gait initiation dynamics in Parkinson’s disease"

### S1: Subject #2 Neural Power Data

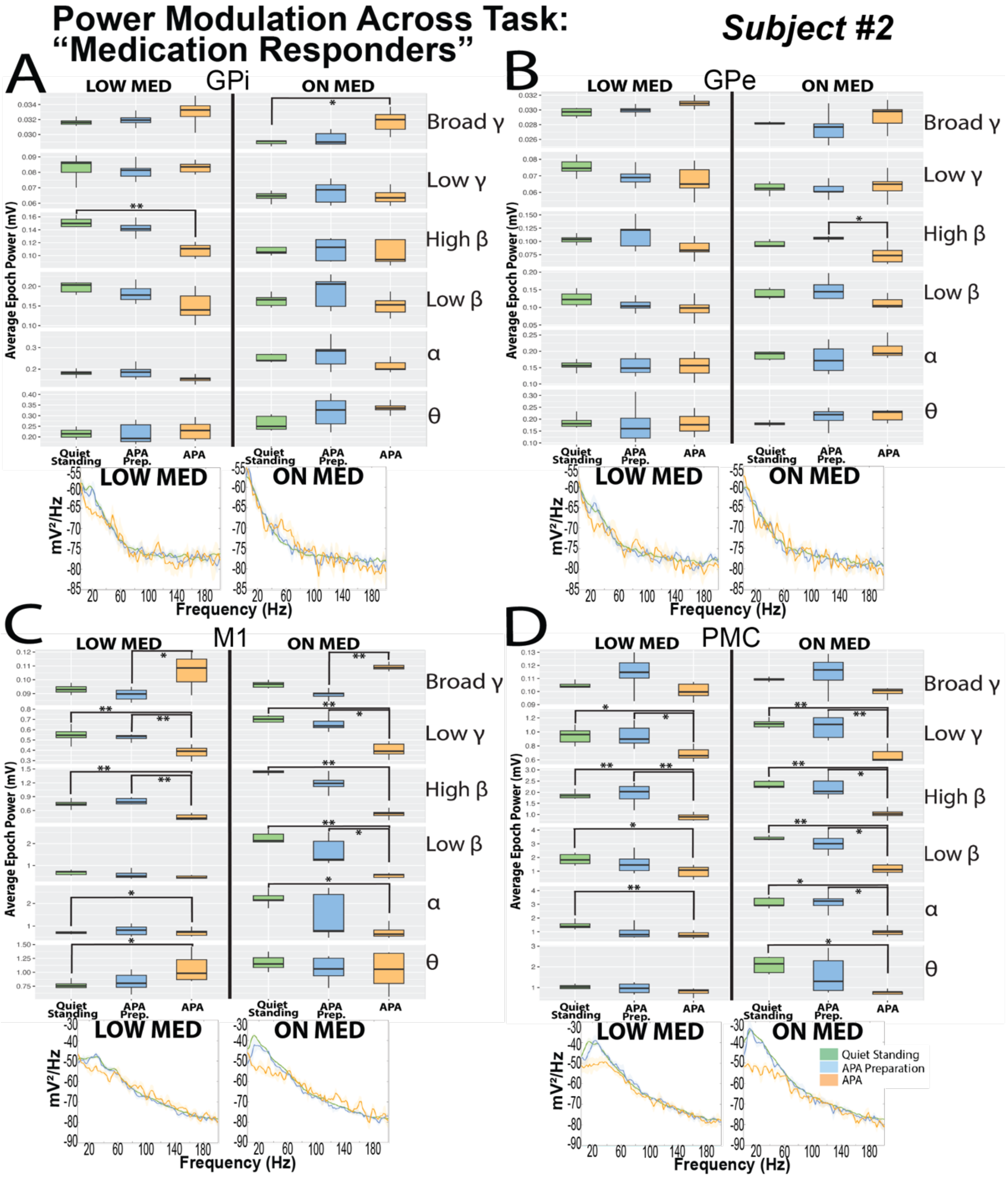

**S2: Subject #4 Neural Power Data**

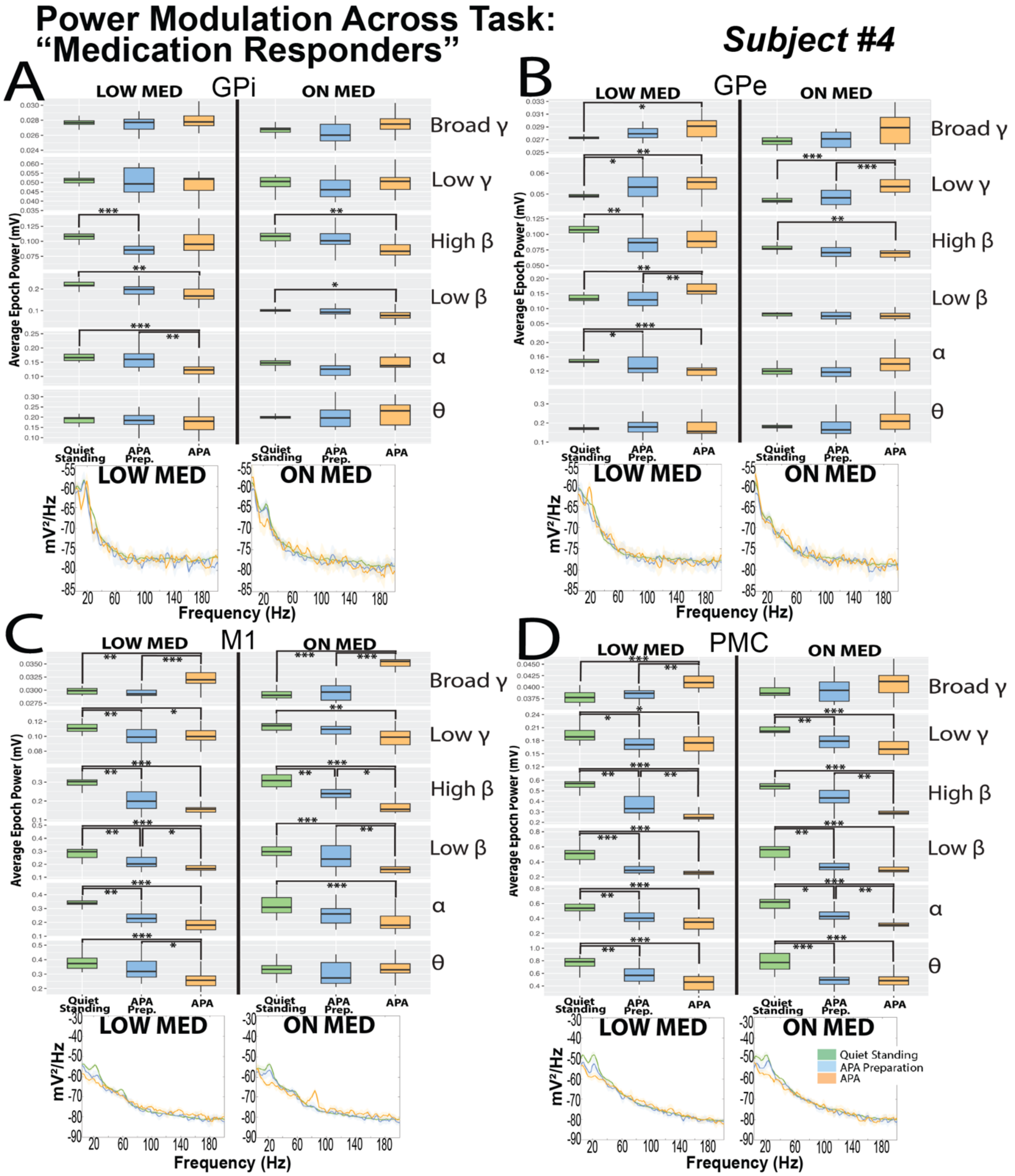

**S3: Subject #1 Neural Power Data**

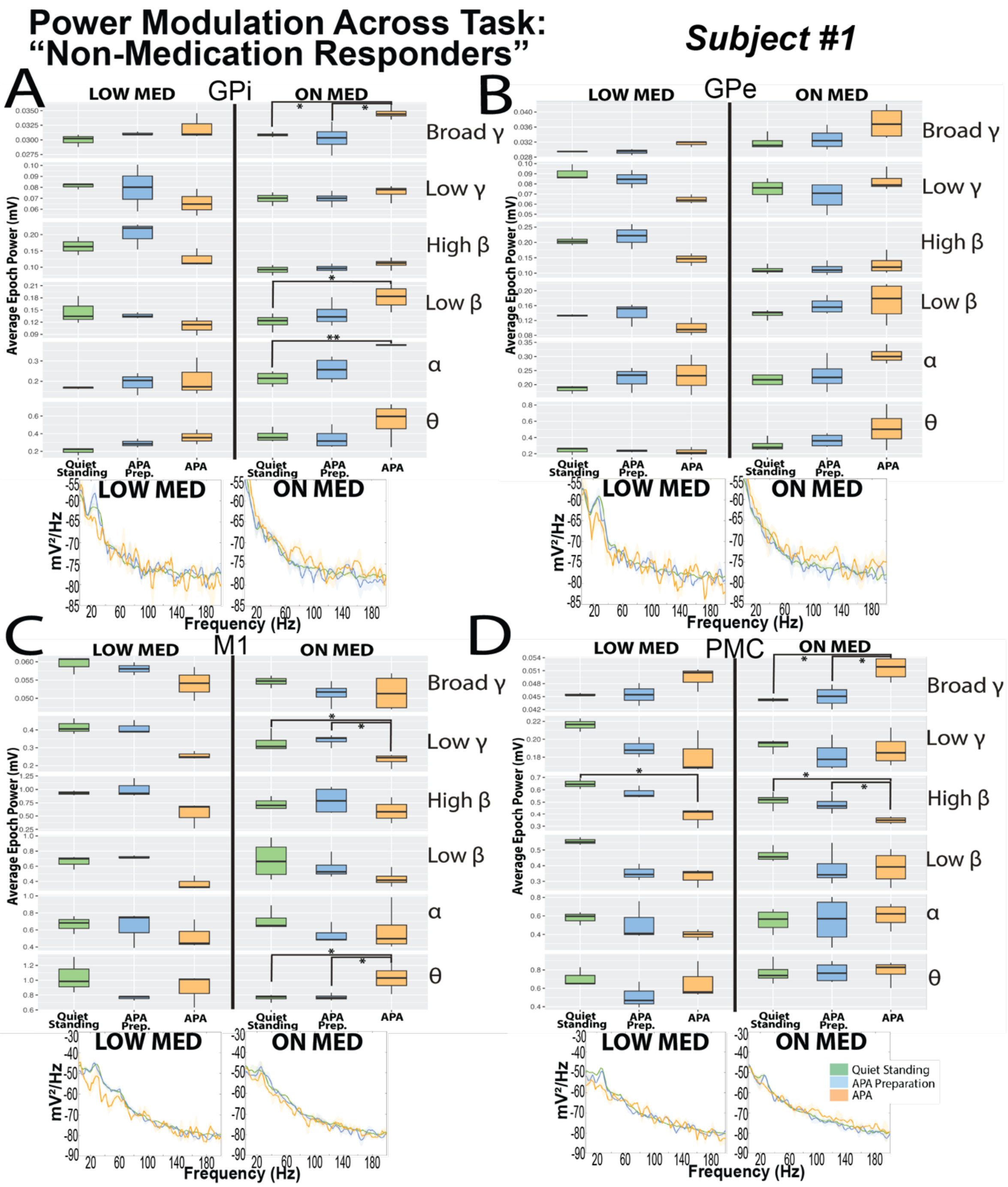

### S4: Subject #3 Neural Power Data

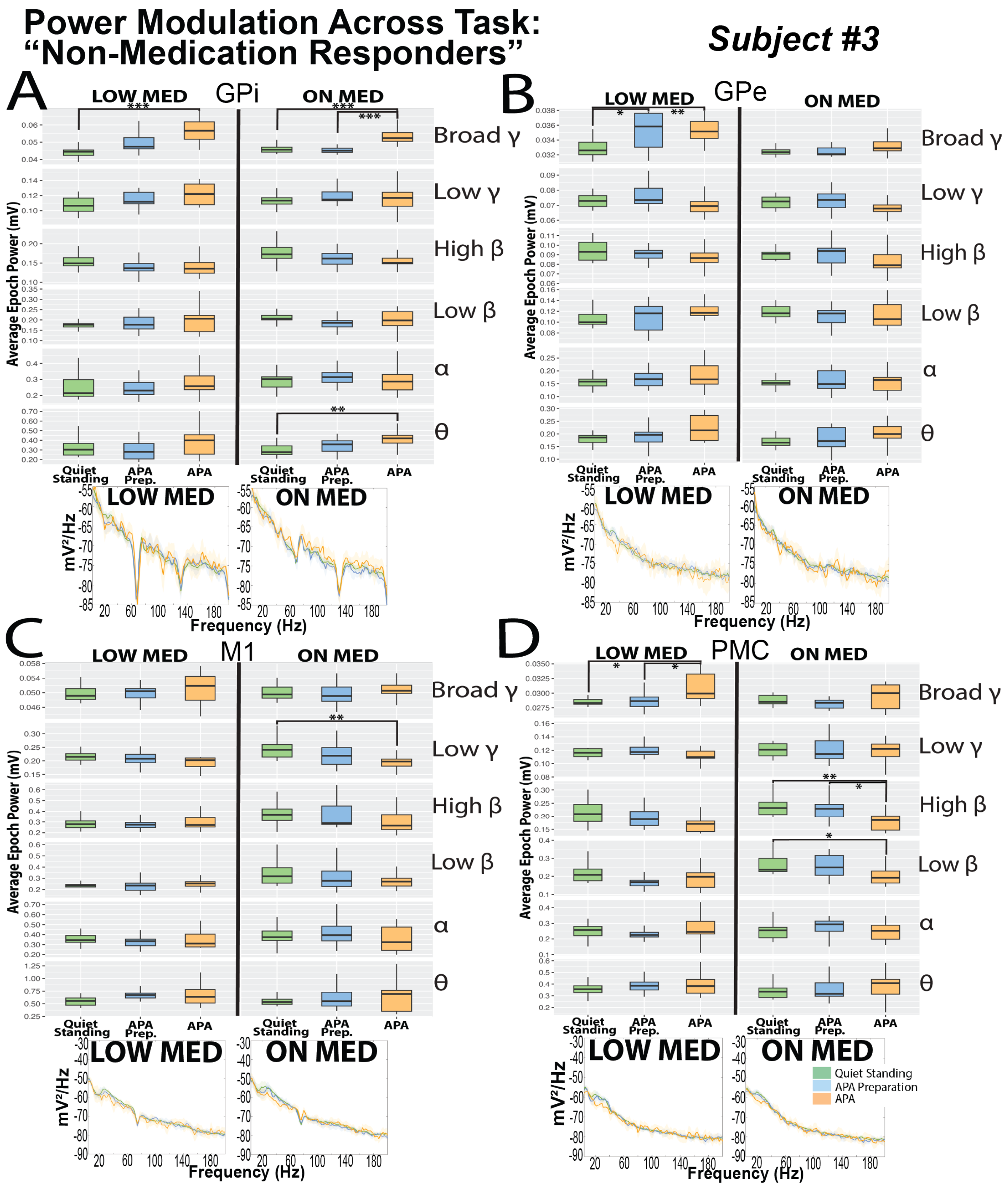

**S5: Subject #5 Neural Power Data**

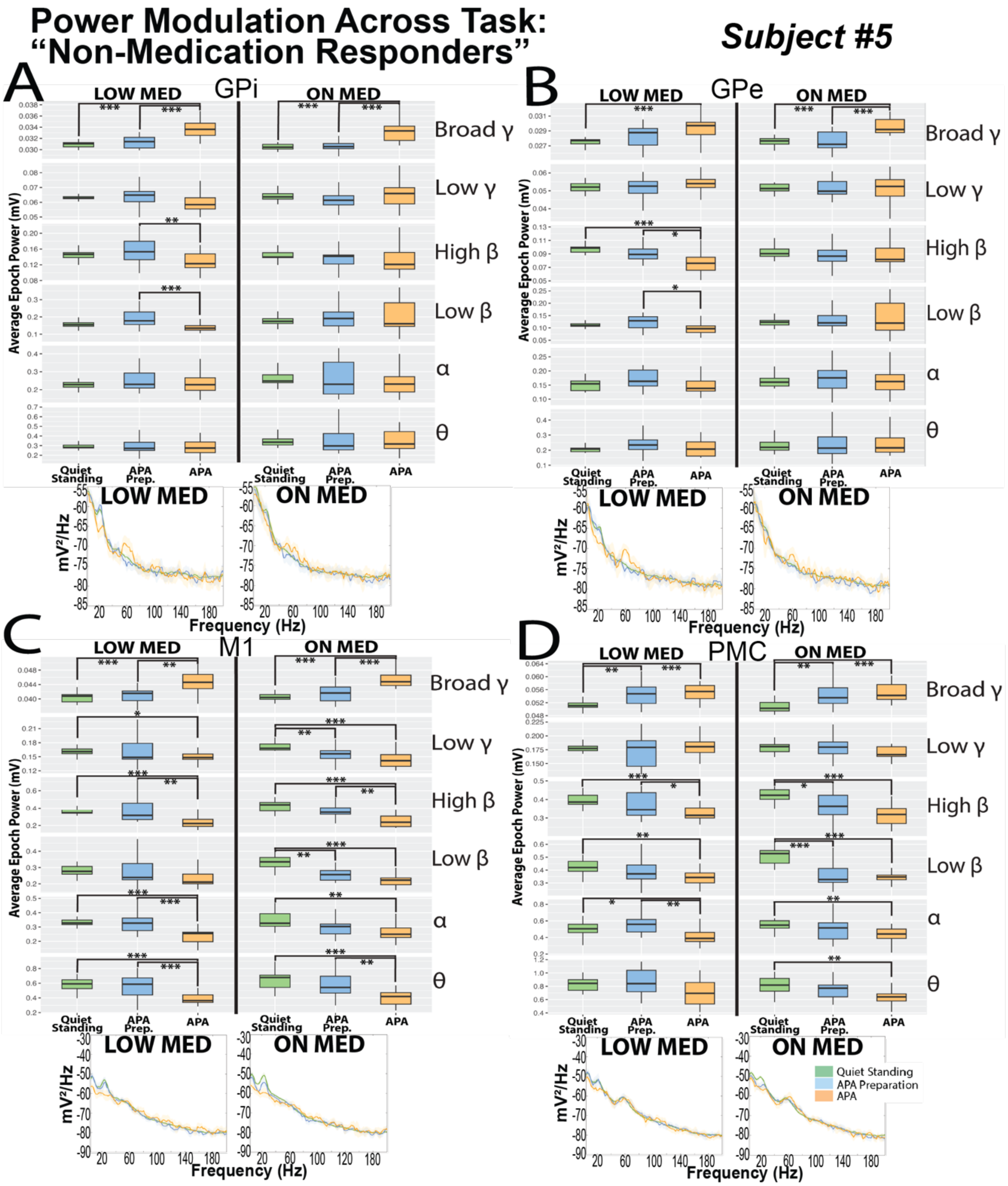


**<u>Supplemental Figures S1-S5:</u>** Individual subject data reflecting power spectra while in the "LOW" and "ON" medication states during the three primary task epochs: Quiet standing (green), APA preparation (blue) and APA (orange). Each panel reflects boxplots demonstrating average power from the "spectrogram" function during each epoch across the canonical frequencies and median PSDs and median absolute deviations for "LOW" and "ON" medication states during the three task epochs: "Quiet standing," "APA preparation" and "APA" at all contacts. GPi (panel A), GPe (panel B), M1 (panel C), PMC (panel D). Significance between epochs is denoted with * $p < 0.05$, ** $p<0.01$, *** $p<0.001$ following Kruskal-Wallis and Dunns testing with multiple corrections. Boxplots created with upper whisker representing 1.5 * interquartile ratio (IQR) past the 3rd quartile and lower whisker representing 1.5 *

the IQR below the 1st quartile. Abbreviations: GPi = globus pallidus internus, GPe = globus pallidus externus, Hz = Hertz; mV = millivolts; APA = anticipatory postural adjustment; θ = theta, α = alpha, β = beta, γ = gamma.

### S6: Group GPi-PMC coherence modulation across the gait initiation task

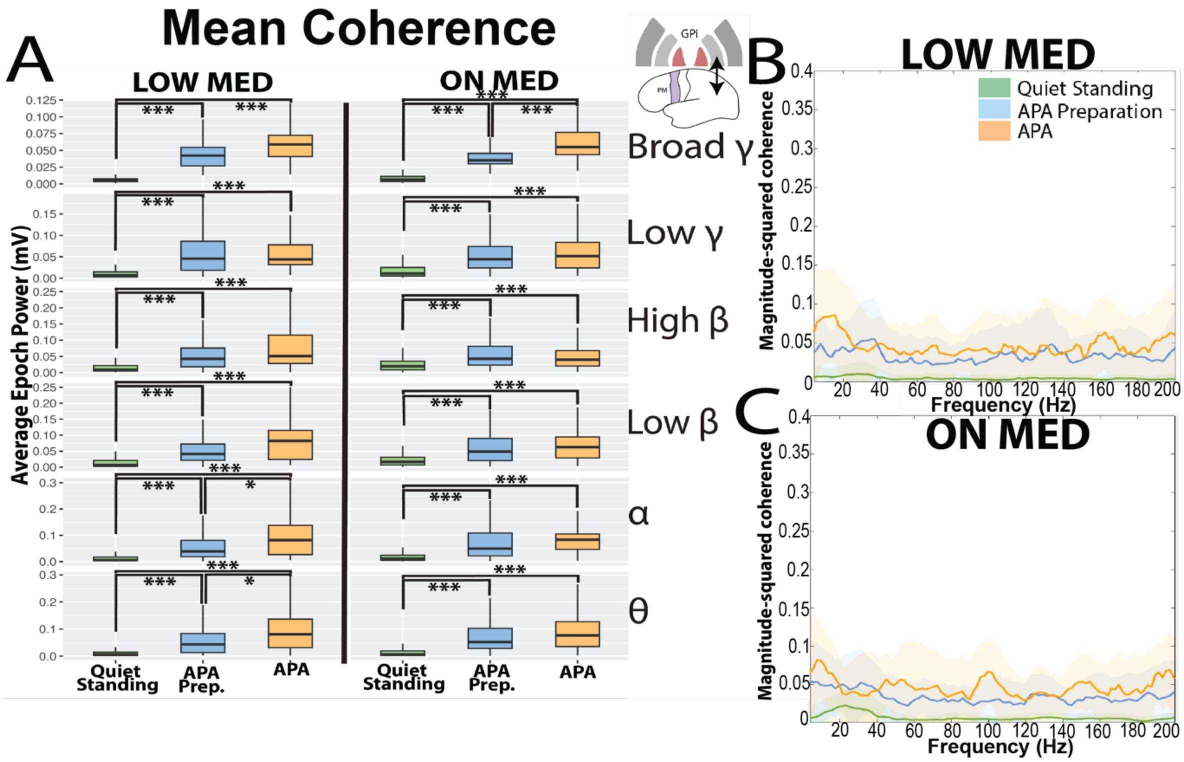


**Supplemental Figure S6:** Grouped subject data across GPi-PMC (panels A-C): B/C reflect median magnitude-squared coherence and median absolute deviations from all subjects in the "LOW" and "ON" medication states during the three primary task epochs: Quiet standing (green), APA preparation (blue) and APA (orange). A: Boxplots represent average magnitude-squared coherence during each epoch across the canonical frequencies. Significance between epochs is denoted with * p< 0.05, ** p<0.01, ***p<0.001 following Kruskal-Wallis testing and multiple corrections. Boxplots created with upper whisker representing 1.5 * the interquartile ratio (IQR) past the 3rd quartile and the lower whisker representing 1.5 * IQR below the 1st quartile. Significant task-related coherence modulation is demonstrated under both medication states and frequencies. Abbreviations: GPi = globus pallidus internus, M1 = primary motor cortex; PMC = premotor cortex, APA = anticipatory postural adjustment; θ = theta, α = alpha, β = beta, γ = gamma.

**S7: Group GPe coherence modulation across the gait initiation task**

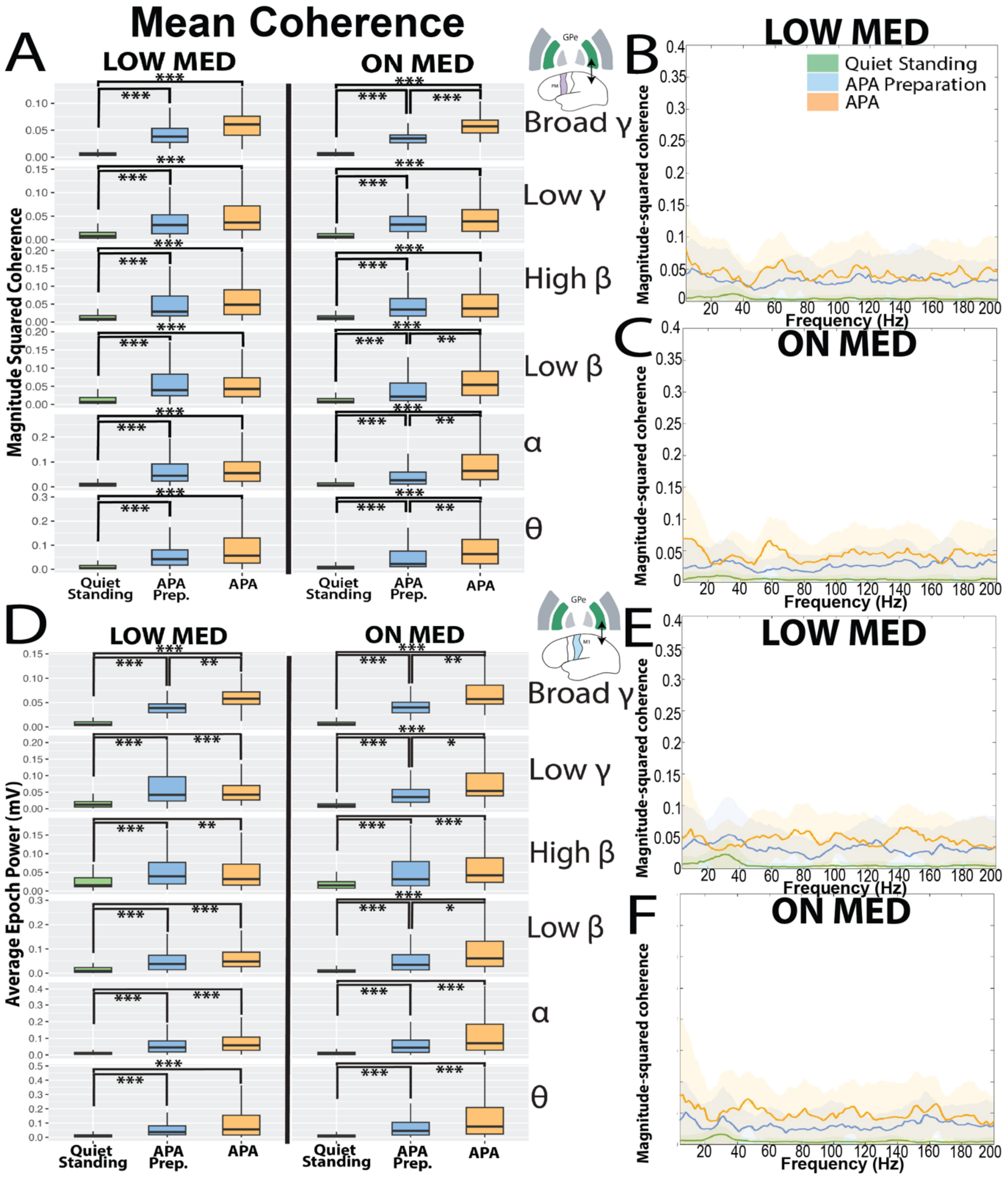


**Supplemental Figure S7:** Grouped subject data across GPe-PMC (panels A-C) and GPe-M1 (panels D-F): B/C and E/F reflect median magnitude-squared coherence and median absolute deviations from all subjects in the "LOW" and "ON" medication states during the three primary task epochs: Quiet standing (green), APA preparation (blue) and APA (orange). A/D: Boxplots represent average magnitude-squared coherence during each epoch across the canonical frequencies. Significance between epochs is denoted with * $p< 0.05$, ** $p<0.01$, ***$p<0.001$ following Kruskal-Wallis testing and multiple corrections. Boxplots created with upper whisker representing 1.5 * the interquartile ratio (IQR) past the 3rd quartile and the lower whisker representing 1.5 * IQR below the 1st quartile. Significant task-related coherence modulation is demonstrated under both medication states and frequencies.

Abbreviations: GPi = globus pallidus internus, M1 = primary motor cortex; PMC = premotor cortex, APA = anticipatory postural adjustment; θ = theta, α = alpha, β = beta, γ = gamma.

### S8: Linear mixed model summary results (Supplementary tables 8A-C)

#### A. Peak APA amplitude metric

| Task Epoch | Normalized data input | AIC | VIFs | Significant predictors (t-statistic) | Random effects: variance (std) |
|---|---|---|---|---|---|
| APA Preparation | Power | 328.7 | 1.0-1.3 | GPi θ (-2.84), GPe θ (2.61), GPe high β (2.00) | Patient ID: 0.30 (0.55)<br>Residual: 1.37 (1.17) |
| **APA** | **Power** | **330.7** | **N/A** | **PMC θ (-2.38)** | **Patient ID: 0.35 (0.59)<br>Residual: 1.45 (1.20)** |
| APA Preparation | Coherence | 322.0 | N/A | GPe-PMC broadband γ (-2.32) | Patient ID: 0.34 (0.58)<br>Residual: 1.47 (1.21) |
| **APA** | **Coherence** | **316.8** | **1.0-4.4** | **GPi-M1 θ (-2.77) & α (3.00), GPe-M1 low β (-2.36)** | **Patient ID: 0.33 (0.57)<br>Residual: 1.33 (1.15)** |

#### B. Change in APA amplitude metric

| Task Epoch | Normalized data input | AIC | VIFs | Significant predictors (t-statistic) | Random effects: variance (std) |
|---|---|---|---|---|---|
| APA Preparation | Power | 226.2 | 1.0-1.1 | GPi low β (-2.36), GPe θ (2.86), GPe high β (2.46) | Patient ID: 0.13 (0.36)<br>Residual: 0.48 (0.69) |
| **APA** | **Power** | **242.0** | **1.0** | **GPi broadband γ (2.47), GPe low γ (-2.64)** | **N/A** |
| APA Preparation | Coherence | 234.6 | 1.0-1.9 | GPe-M1 α (-2.62), high β (2.22), low γ (-2.22), broadband γ (-2.39) | N/A |
| **APA** | **Coherence** | **220.9** | **4.4** | **GPi-M1 θ (-3.50) & α (2.79)** | **Patient ID: 0.21 (0.46)<br>Residual: 0.48 (0.69)** |

#### C. Time to peak APA amplitude

| Task Epoch | Normalized data input | AIC | VIFs | Significant predictors (t-statistic) | Random effects: variance (std) |
|---|---|---|---|---|---|
| APA Preparation | Power | -221.9 | 1.2-1.4 | GPi high β (-2.51), M1 θ (-2.75), M1 low β (2.55) | N/A |
| **APA** | **Power** | **-215.2** | **N/A** | **N/A** | **N/A** |
| APA Preparation | Coherence | -238.7 | 1.1 | GPe-M1 α (-2.71), low β (2.33), broadband γ (4.61), M1-PMC broadband γ (2.24) | N/A |
| **APA** | **Coherence** | **-221.1** | **1.0-1.1** | **GPi-M1 θ (-2.10), GPe-M1 α (-3.28), low β (2.78), M1-PMC broadband γ (-2.15)** | **N/A** |

**Supplemental Table S8:** Summary statistics from the LMMs for each APA metric from the three primary task epochs, which were normalized to quiet standing. AIC = Akaike Information Criterion; VIF = variance inflation factor. Random effects presented as the variance (standard deviation); if "N/A" this indicates the optimized model did not include this as a significant predictor. APA = anticipatory postural adjustment, GPi = globus pallidus internus, GPe = globus pallidus externus, M1 = primary motor cortex, PMC = premotor cortex.
